# KZFP-mediated variable DNA methylation of a primate-specific transposon is linked to type I diabetes in humans

**DOI:** 10.1101/2025.04.22.25326054

**Authors:** Maria Derakhshan, Prachand Issarapu, Andrew M Prentice, Julius A Kuebel, Sophie E Moore, Robin M Bernstein, Abdul K Sesay, Mariama Jammeh, Robert A Waterland, Garrett Hellenthal, Michael Imbeault, Matt J Silver

## Abstract

Variable DNA methylation states during early embryonic development overlap with mechanisms of transposon silencing by Krüppel-associated box zinc finger proteins (KZFPs). We investigated the influence of genetic variation in KZFPs and their transposon targets and identified a variably methylated region (VMR) proximal to the human *LY6S-AS1* gene linked to an intronic MER11C retrotransposon targeted by the primate-specific KZFP ZNF808. Mendelian randomisation analysis supported a causal link between VMR methylation and type 1 diabetes risk, and H1 stem cells with inactivated *ZNF808* showed marked, transient upregulation of the *LY6S-AS1* transcript during the early stages of pancreatic development. Loss of function genetic mutations in *ZNF808* have previously been linked to pancreatic agenesis and neonatal diabetes in humans, and the VMR was previously associated with levels of insulin secretion in Gambian children. Together this evidence points to a link between KZFP-mediated DNA methylation of *LY6S-AS1*, and pancreatic development and function at this locus.

## Introduction

Transposable elements can act as a novel source of gene regulation, yet they have the potential to destabilise the genome and their repression is fundamental to genome integrity^1,2^. In vertebrates, KZFPs epigenetically silence transposable elements through recruitment of KAP1/TRIM28, SETDB1 and DNA methyltransferases, leading to the establishment of trimethylated histone H3 Lys9 (H3K9me3) and DNA methylation (DNAm) marks during early embryonic development^3–5^. In mice, retrotransposons are influenced by a diverse set of *trans*-acting KZFPs with epigenetic silencing capabilities that can result in inter-individual differences in DNA methylation states^6,7^. In isogenic mice, some of these variably DNA methylated loci are described as ‘metastable epialleles’ (MEs) characterised by inter-individual variation that is consistent across tissues, indicative of differential establishment in the early embryo^8^. A notable example is the agouti locus in Agouti viable yellow (*A*^*vy*^) mice where a variably methylated intracisternal A-particle (IAP) retrotransposon upstream of the agouti gene influences gene expression and a range of phenotypes including coat colour, obesity and diabetes^9^.

Putative MEs have also been identified in humans and are associated with obesity, thyroid function and several cancers^10^. We recently identified a set of hypervariable CpGs (hvCpGs) with variable methylation states consistently observed across 19 tissues and 8 ethnicities^11^. hvCpG methylation covaried across embryonic tissues derived from different germ layers, and hvCpGs were enriched for proximity to KZFP binding sites and ERV1 and ERVK retrotransposons, positioning these loci as potential human counterparts to murine MEs^10,11^.

Here, we report the identification of a VMR comprising multiple hvCpGs overlapping the promoter of the *LY6S-AS1* long non-coding RNA gene. Methylation at this locus is associated with genetic variation in the primate-specific KZFP ZNF808 and its binding site in a MER11C retrotransposon located in the *LY6S-AS1* first intron. Mendelian randomisation and functional analysis of H1 stem cells suggests a causal link to pancreatic development and function at this locus. The VMR has previously been associated with levels of insulin secretion in Gambian children^12^, and loss of function genetic mutations in *ZNF808* are linked to pancreatic agenesis and neonatal diabetes in humans through the control of MER11 elements^13^, suggesting a role for KZFP-mediated epigenetic silencing of *LY6S-AS1* in pancreatic development and function.

## Results

### KZFP-mediated silencing of retrotransposons is linked to hypervariable methylation in humans

Investigation of the influence of genetic variation on DNAm through methylation quantitative trait loci (mQTL) mapping can help elucidate the effects of transcription factors on gene regulation^14^. We examined the role of KZFP-mediated epigenetic silencing at hvCpGs by analysing *trans-*mQTL in KZFP genes and links to their retrotransposon targets (Figure 1A).

**Figure 1.**
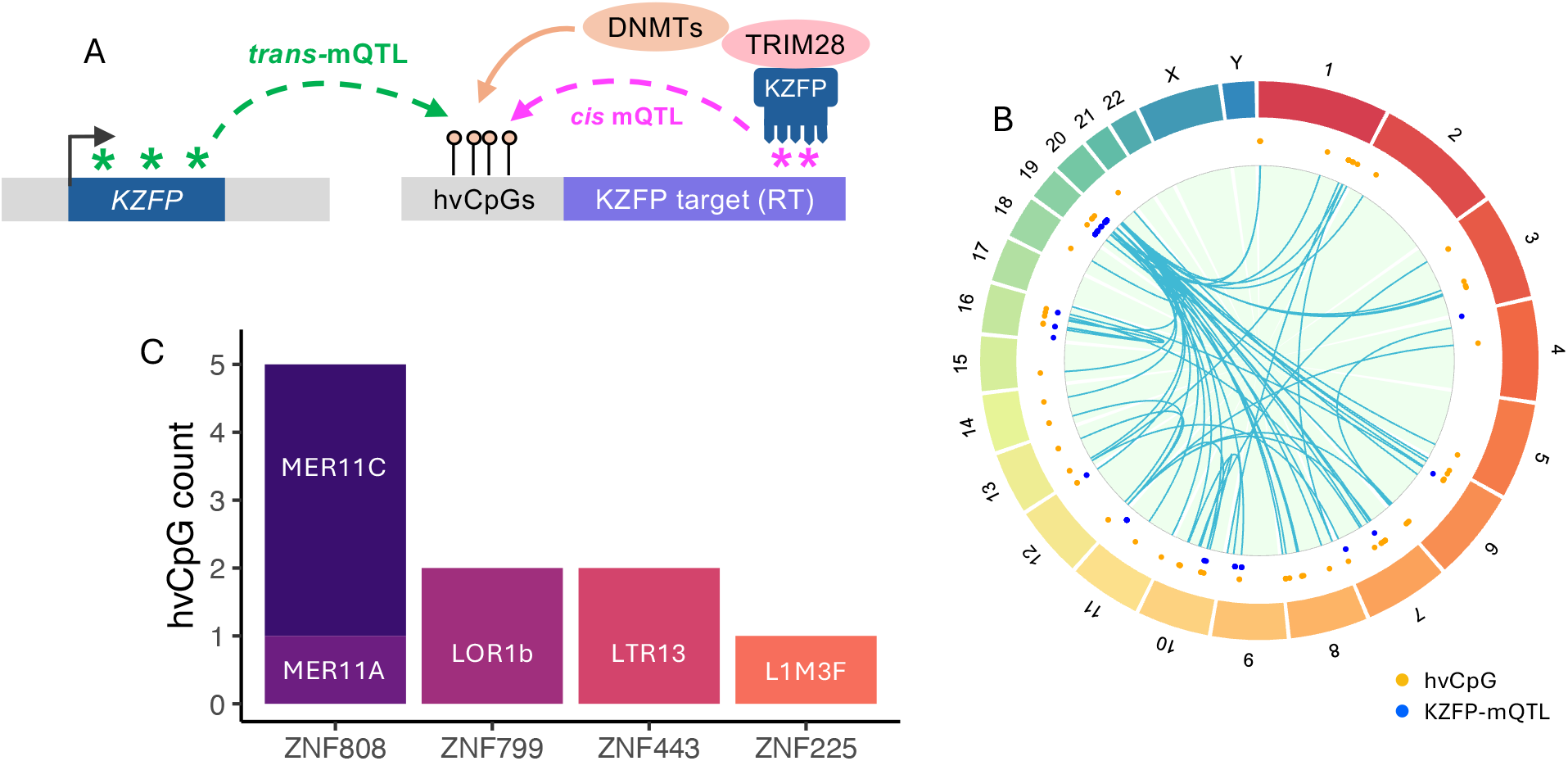
KZFP-mediated TE silencing is linked to hypervariable methylation in humans. **A:** Schematic representation of a transcribed KZFP binding to its target RT and recruiting silencing co-factors including TRIM28 and DNMTs that methylate the RT and a neighbouring cluster of hvCpGs. Genetic variation in the KZFP gene (*trans-*mQTL) can influence hvCpG methylation by affecting KZFP expression, binding at the target RT and/or recruitment of cofactors. Genetic variation at the target RT binding site (*cis-*mQTL) can influence hvCpG methylation by affecting KZFP binding. **B:** Associations between 137 hvCpGs (orange points) and 105 KZFP mQTL (blue points). Chromosomes are labelled on the outer circle. **C:** Distribution of hvCpGs with KZFP *trans*-mQTL, where the KZFP target RT is within 3kb of the hvCpG and a TRIM28 ChIP-exo site. *ZNF808* mQTL are associated with 5 hvCpGs, proximal to two RTs (MER11A and MER11C) targeted by ZNF808. ZNF799, ZNF443 and ZNF225 target single RTs LOR1b (mQTL associated with 2 hvCpGs), LTR13 (2 hvCpGs) and L1M3F (1 hvCpG) respectively. See Supplementary Table 2 for complete data on all *trans-*mQTL associations. mQTL: methylation quantitative trait loci; RT: retrotransposon; hvCpG: hypervariable CpG; DNMTs: DNA methyltransferases.

Since hvCpGs show robust, hypervariable methylation across multiple tissues and ethnicities^11^, we analysed data on blood mQTL from a large meta-analysis of 32,851 individuals of predominantly European ancestry conducted by the Genetics of DNA Methylation Consortium (GoDMC)^14^. *Trans*-mQTL for hvCpGs (GoDMC threshold p < 1 x 10^-14^) were enriched within 366 human protein-coding KZFP genes^1^ (10.8% KZFP *trans*-mQTL for hvCpGs compared to 4.6% for the Illumina methylation array background with *trans*-mQTL; odds ratio (OR) = 2.3; Fisher’s exact test (FET) p = 5.5 x 10^-12^; Methods). In total, we identified 142 associations between 137 hvCpGs (clustering into 92 VMRs) and 105 *trans*-mQTL located in 65 KZFP genes (Figure 1B; Supplementary Table 1).

KZFPs recruit TRIM28 as part of their silencing machinery^4^ and since methylation at hvCpGs is established early in development, we next examined the proximity of hvCpGs to TRIM28 recruitment sites identified by chromatin immunoprecipitation with exonuclease digestion (ChIP-exo) in human embryonic stem cells (H1-ESCs)^1^. hvCpGs were more likely to be located within 3kb of a TRIM28 ChIP-exo peak, compared to other CpGs on the array background (10.1% hvCpGs compared to 4.2% array background; OR = 2.6, FET p = 1.3 x 10^-60^). TRIM28 recruits SETDB1 which deposits repressive H3K9me3 marks associated with heterochromatin in the early embryo^15^. Accordingly, hvCpGs were enriched for overlapping solo H3K9me3 modifications (OR = 1.5, FET p = 2.4 x 10^-9^) and for dual/tri histone modifications including H3K9me3 (OR = 1.3-2.0, FET p = 0.002-3.7×10^-14^) in H1-ESCs compared to array background (Supplementary Figure 1).

Finally, we identified a subset of hvCpGs with KZFP *trans*-mQTL that were proximal (<3kb) to the retrotransposon targeted by that KZFP, and to a TRIM28 ChIP-exo peak in H1 stem cells. This resulted in a total of 10 hvCpGs falling into 6 clusters that were associated with *trans-*mQTL in 4 distinct KZFPs, targeting 5 distinct retrotransposons (Figure 1C; Supplementary Table 2).

### An hvCpG cluster upstream of *LY6S-AS1* is associated with genetic variation at *ZNF808* in *trans* and its proximal MER11C retrotransposon target in *cis*

5 of the 10 hvCpGs described above mapped to *trans-*mQTL in the *ZNF808* gene (Figure 1C). Of these, 4 hvCpGs form a cluster overlapping the promoter of the long non-coding RNA gene *LY6S-AS1* (hereafter ‘LY6S-VMR’; Figure 2A; Supplementary Table 2). ZNF808 primarily targets subfamilies of MER11 endogenous retroviral elements^13^. Accordingly, *LY6S-AS1* contains a MER11C element in the first intron, located 1,500bp downstream of the LY6S-VMR (Figure 2A). Data from ChIP-exo of TRIM28 in H1-ESCs and KZFPs in HEK293T cells^1^ confirms binding of TRIM28, ZNF808 and ZNF525 at this element. Furthermore, one of the 4 hvCpGs in the LY6S-VMR (Illumina ID cg077702222) shows evidence of systemic interindividual variation (SIV), another (cg04612566) demonstrates epigenetic supersimilarity^16^ (Supplementary Table 3), and the VMR is 7.6kb upstream of a correlated region of SIV^17^. These properties are indicative of DNA methylation establishment in the early embryo at the LY6S-VMR, supporting a link to ZNF808-mediated silencing at this point in early development^10,11^.

**Figure 2.**
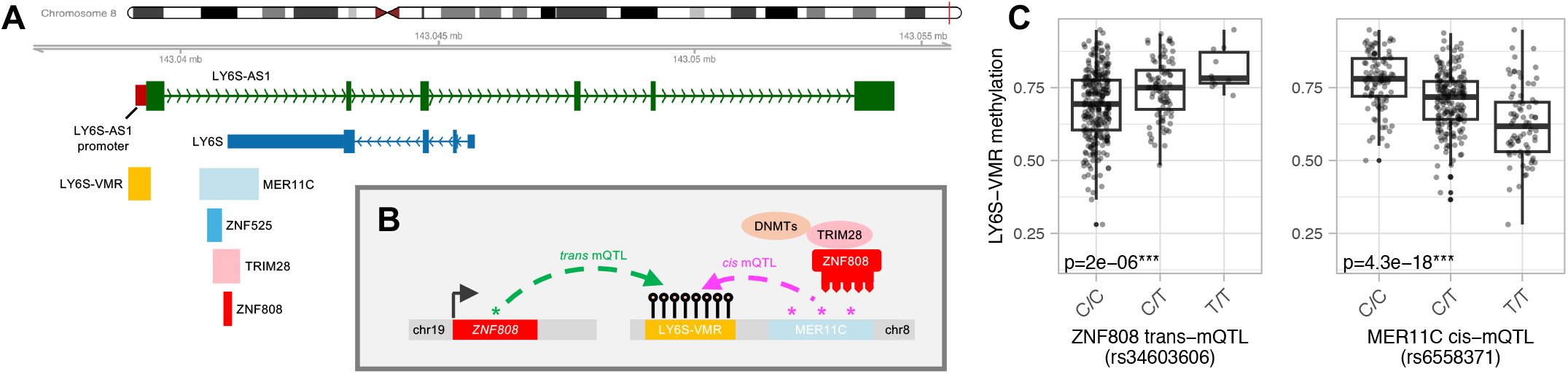
The LY6S-VMR is associated with genetic variation at *ZNF808* in *trans* and its proximal MER11C retrotransposon target in *cis*. **A:** Schematic of the *LY6S-AS1* long non-coding RNA gene on chromosome 8 with exons marked as green blocks. The LY6S*-*VMR overlaps the *LY6S-AS1* promoter. A MER11C element and ZNF808, ZNF525 and TRIM28 H1-ESC ChIP-exo peaks are located in the *LY6S-AS1* first intron. **B:** Methylation at all 8 CpGs in the LY6S-VMR is associated with genetic variation at the KZFP ZNF808 (mQTL in *trans* on chromosome 19) and its proximal MER11C RT target (mQTL in *cis* on chromosome 8), in mQTL data from GoDMC. **C:** Significant associations at a single LY6S-VMR CpG (chr8: 144120111) with mQTL in *trans* (*ZNF808*, left) and in *cis* (MER11C, right) in genotype and blood DNA methylation data from the Gambian ENID cohort (n=359). Associations across all CpGs in the VMR are shown in Supplementary Figure 3. ‘***’ indicates significant association accounting for multiple tests (Methods).

To further investigate the influence of ZNF808-mediated MER11C silencing at the LY6S-VMR, we first extended our analysis of GoDMC *trans*-mQTL to the 8 CpGs falling within the LY6S-VMR on the HM450k array (Supplementary Table 3). All 8 CpGs had highly significant associations with one or more mQTL distributed across *ZNF808* exons and introns (maximum p = 8×10^-181^; Supplementary Table 4). The mQTL were in strong LD (r^2^>0.8) with 116 SNPs distributed across the gene and neighbouring regions (Supplementary Figure 2).

Next, we replicated the association of *ZNF808 trans*-mQTL with LY6S-VMR CpGs in the GoDMC dataset using our own data comprising methyl-seq and imputed genotype data from blood samples taken from the Gambian ‘ENID’ cohort of 2 year-old infants (Methods)^18,19^. We tested associations between 78 *ZNF808* SNPs and 13 CpGs falling within the LY6S-VMR in 359 individuals with available data (Supplementary Table 3; Methods). After correction for multiple testing, one SNP (rs34603606) was significantly associated with methylation at 3 LY6S-VMR CpGs (maximum p=2.3×10^-5^; Supplementary Tables 5 and 6; Figure 2C; Supplementary Figure 3A), replicating in Africans the observed link between *ZNF808* and the LY6S-VMR in GoDMC data from Europeans.

Finally, we investigated the impact of genetic variation in the proximal MER11C element on LY6S-VMR methylation by analysing mQTL in *cis*. All 8 LY6S-VMR CpGs on the 450k array were associated with one or more *cis*-mQTL in the GoDMC meta-analysis after LD clumping^14^ (maximum p = 1.8×10^-33^; Supplementary Table 7). Analysis of unclumped *cis*-mQTL revealed that one SNP (rs6558371) in the ZNF808 MER11C ChIP-exo peak was associated with all 8 LY6S-VMR CpGs (all p=0; Supplementary Table 8). This SNP was also significantly associated with all 13 LY6S-VMR CpGs in the Gambian methyl-seq data (maximum p = 2.0 x 10^-5^; Figure 2C; Supplementary Figure 3B; Supplementary Table 6).

### Nanopore sequencing reveals relationship between MER11C silencing and LY6S-AS1 VMR methylation

Next, we characterised regional methylation at the LY6S-VMR and its proximal MER11C in high resolution using Nanopore long-read sequencing data generated from n=88 blood DNA samples from an independent cohort of 1-year olds from the Gambian ‘HERO-G’ cohort (Methods). Analysis of these data showed that the LY6S-AS1 proximal MER11C is mostly methylated, but that variable methylation at the LY6S-VMR depends on varying patterns of methylation decay from the MER11C element (Figures 3A and 3B; Supplementary Figure 4). Read-level data from a single individual that was hypomethylated at the VMR (Figure 3C, purple box) showed an increased proportion of DNA molecules with loss of methylation across the VMR, compared to an individual that was hypermethylated (red box). This accords with our previous observation of inter-cellular methylation variegation effects at putative human MEs, measured by whole-genome bisulfite-seq^20^.

**Figure 3.**
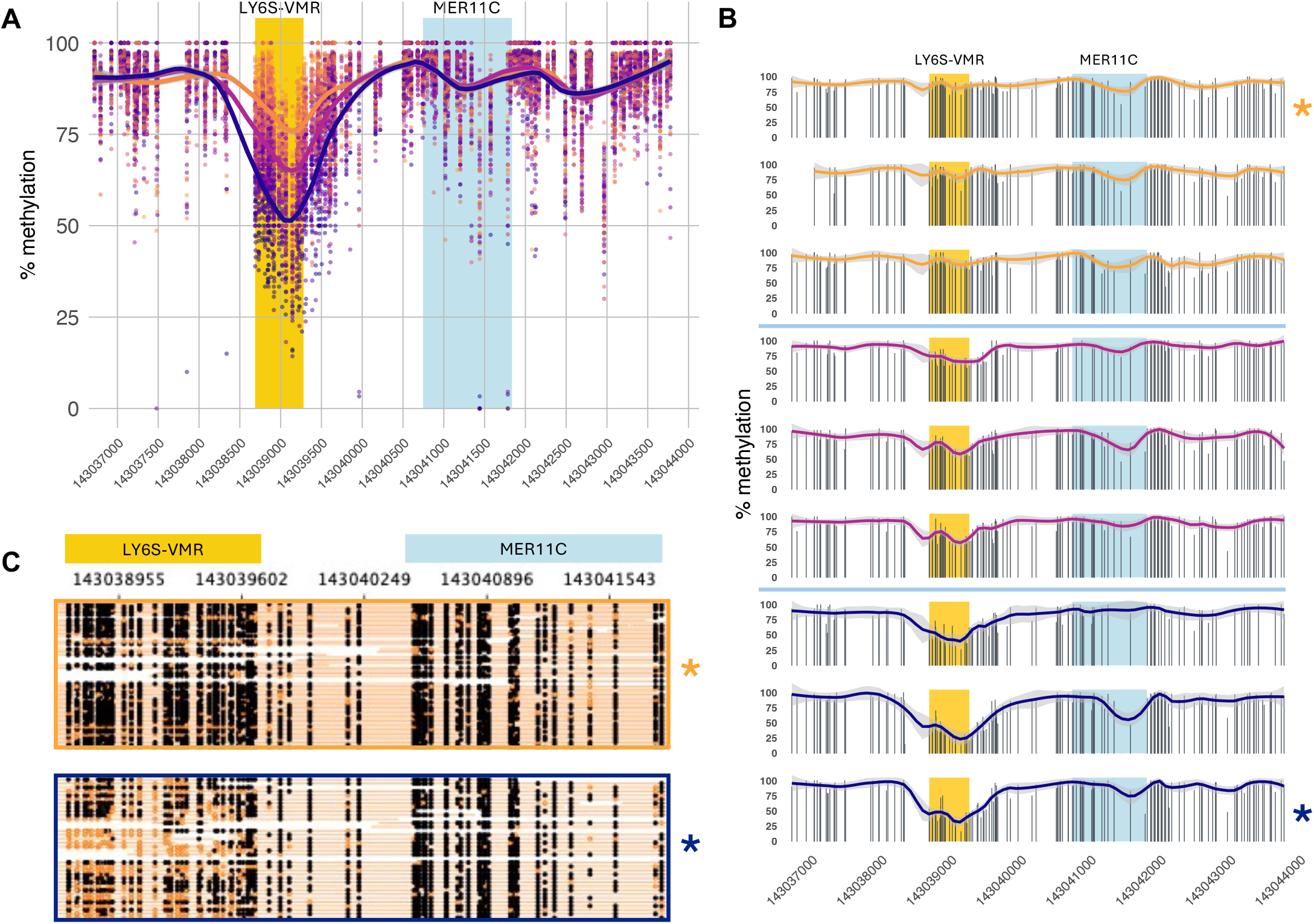
High-resolution characterisation of the LY6S-VMR - MER11C region with Nanopore methyl-seq data. Blood methylation data from 1 year-old infants from the Gambian HERO-G cohort. **A:** CpG methylation values for 88 samples (points coloured by sample ID) with at least 75% of CpGs across the region covered at a minimum depth of 20x. Loess curves summarise regional methylation values for samples in the top, middle and bottom tertiles of mean methylation across the LY6S-VMR. X-labels are CpG coordinates. **B:** Individual-level plots for 9 samples (from **A**) that have the highest (top 3 plots), intermediate (middle 3) and lowest (bottom 3) mean methylation across the LY6-VMR. Black bars indicate methylation values for CpGs covered at more than 20x. 95% confidence intervals for loess curve fits are shown in light grey. **C:** Read-level plots for two samples from **B** (marked with asterisks), that have the highest (top) and lowest (bottom) mean methylation across the LY6S-VMR. Orange horizontal lines indicate individual reads. Methylated CpGs are filled black circles. Unmethylated CpGs are unfilled orange circles. All coordinates hg38.

### The LY6S-VMR overlaps a DMR associated with insulin secretion in Gambians, with evidence of a causal link to type 1 diabetes risk in Europeans

Homozygous loss of function mutations in *ZNF808* have been linked to neonatal diabetes driven by pancreatic agenesis^13^. Notably, 7 out of 8 LY6S-VMR overlap a differentially methylated region (hereafter ‘insDMR’) associated with insulin secretion in an independent cohort of Gambian children^12^ (Supplementary Table 3). These findings prompted us to investigate whether methylation at the insDMR may be causally linked to type 1 diabetes (T1D), type 2 diabetes (T2D) or pancreas volume (PV), using a two-sample Mendelian randomization (2SMR) framework^21^ (Supplementary Figure 5).

We used 11 GoDMC mQTL in *cis* and in *trans* as genetic instruments for the seven Illumina 450K CpGs in the insDMR (Supplementary Tables 3 and 9) and extracted summary level data for these SNPs from T1D, T2D and PV GWAS in Europeans that were available in the Open GWAS database (https://gwas.mrcieu.ac.uk/datasets; Supplementary Table 10, Methods).

After accounting for multiple tests, 2SMR analysis of T1D GWAS provided evidence of a causal link between insDMR methylation and increased risk of T1D at multiple CpGs in two out of three analysed GWAS (median [IQR] p = 0.002 [8×10^-4^,0.02]; Figure 4; Supplementary Table 11). We found no evidence of weak instrument bias or pleiotropic SNP effects (Supplementary Table 11; Methods) and a test for directionality suggested a causal effect of insDMR methylation on T1D risk (Supplementary Table 11; Methods). We found no evidence for causal associations between the iDMR and T2D or PV (Supplementary Figures 6 & 7; Supplementary Tables 12 & 13).

**Figure 4.**
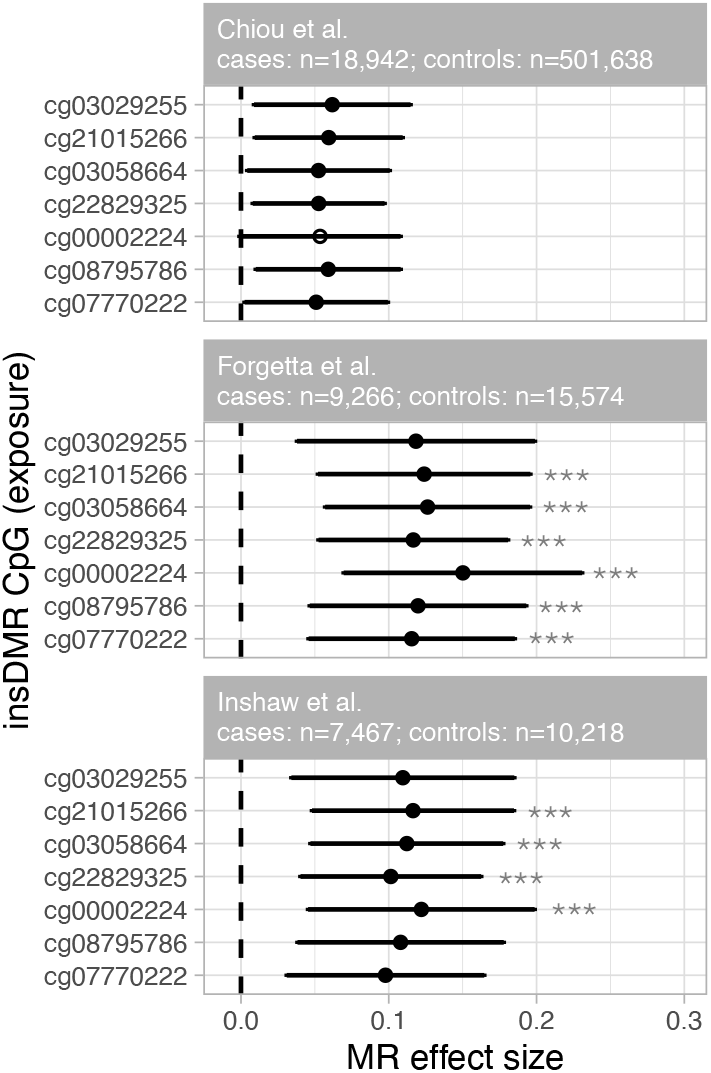
Two-sample Mendelian randomisation (2SMR) supporting a causal link between LY6S-VMR methylation and T1D risk. Forest plots show 2SMR effect estimates (x-axis) for CpGs in the insDMR (y-axis) for data from three GWAS. Points show the mean causal effect of each CpG on T1D risk. Horizontal lines are 95% confidence intervals. ‘***’ indicates significant association accounting for multiple tests. See Supplementary Table 10 for GWAS details and Supplementary Table 11 for full results.

Together, these results suggest a potential causal link between LY6S-AS1 VMR methylation and T1D risk.

### Functional impact of *ZNF808* inactivation on regulation and *LYS6-AS1* expression during endoderm differentiation

De Franco *et al*. highlighted the importance of ZNF808 for the maintenance of H3K9me3-dependent heterochromatin at MER11 elements and for the appropriate specification of pancreatic cells during endoderm differentiation^13^. We reanalysed transcriptomic and epigenetic data from this published dataset to probe for evidence of MER11-mediated regulatory activity at the *LY6S-AS1* locus. Analysis of RNA-seq data showed that *LY6S-AS1* has low expression in wildtype H1 stem cells but is dramatically and transiently upregulated at the definitive endoderm stage (Figure 5A). In *ZNF808* KO cells there is significant upregulation of the LY6S-AS1 transcript in the stem cell state compared to wildtype cells (S0, 2.8 fold change (FC), false discovery rate (FDR) 1×10^-5^), definitive endoderm (S1, FC 3.0, FDR 1×10^-23^) and primitive gut tube stage (S2, FC 1.6, FDR 1×10^-4^). Furthermore, at the definitive endoderm stage we observed complete loss of H3K9me3 epigenetic marks at the *LY6S-AS1* associated MER11C element with a moderate increase in H3K27ac levels both at the *LY6S-AS1* promoter and in the region surrounding the MER11C (Figure 5B). While De Franco *et al*. did not quantify DNA methylation, these findings point to a functional role for ZNF808-mediated repression of MER11C at this locus.

**Figure 5.**
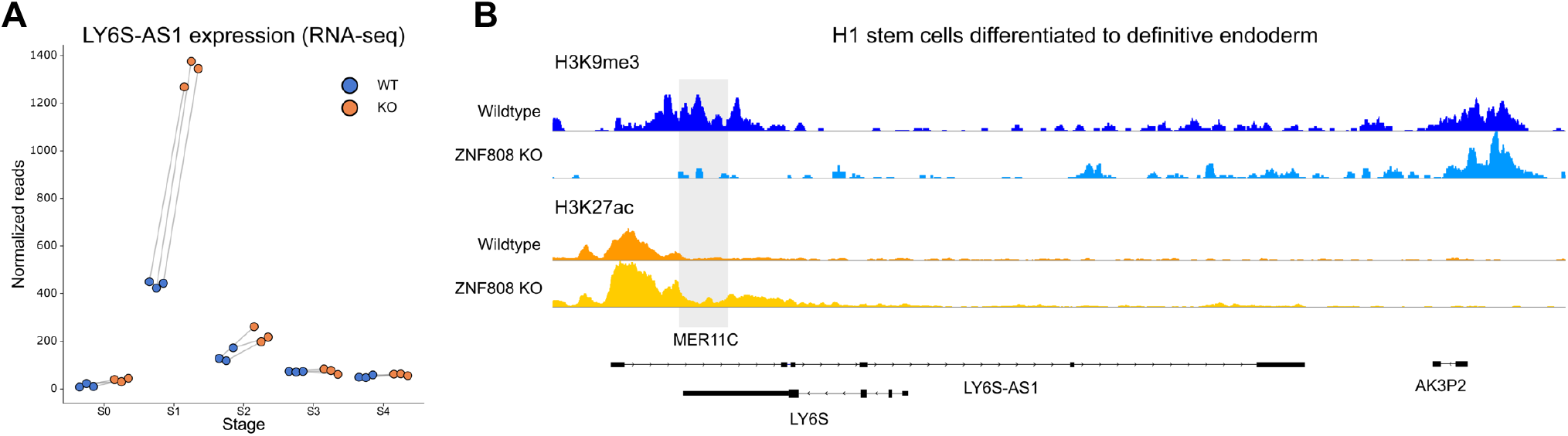
Impact of *ZNF808* inactivation on regulation and expression of LYS6-AS1 during endoderm differentiation. **A:** Analysis of RNA-seq data in differentiating endoderm shows significant upregulation of *LY6S-AS1* in *ZNF808* KO cells in the stem cell state (S0), definitive endoderm (S1) and primitive gut tube stage (S2). **B:** H3K9me3 epigenetic marks (top) are lost at at the definitive endoderm stage in *ZNF808* KO cells at the LY6S-AS1 associated MER11C element. H3K27ac levels (bottom) at the LY6S-AS1 promoter and in the region surrounding the MER11C show a moderate increase.

## Discussion

Retrotransposons are linked to disease in humans^22^ and KZFPs are instrumental in their epigenetic silencing during embryonic development through deposition of H3K9me3 and DNA methylation^23^. We assessed the influence of KZFP-mediated silencing of retrotransposons on DNA methylation at proximal, variably methylated CpGs in humans. Through analysis of mQTL in *cis* and in *trans*, and through interrogation of long-read sequencing data, we found evidence that methylation at a VMR in the promoter of the lncRNA gene *LY6S-AS1* is linked to silencing of its adjacent MER11C by the primate-specific KZFP ZNF808. Evidence of systemic interindividual variation^20^ and epigenetic supersimilarity^16^ at LY6S-VMR CpGs suggests that LY6S-VMR methylation is established during early embryonic development.

Coupled with these findings, our observation that ZNF808 inactivation directly influences *LY6S-AS1* expression during early pancreatic development suggests this gene may be a key component of the pathway linking *ZNF808* inactivation to pancreatic agenesis and neonatal diabetes through epigenetic silencing of its intronic MER11C^13^. Moreover, methylation variation at the LY6S-VMR is associated with insulin secretion in Gambian children^12^, and we provide evidence of a causal link between LY6S-VMR methylation and T1D risk supporting a potential influence on phenotype.

Other factors driving variable methylation states at the LY6S-VMR remain to be elucidated. While the *cis* and *trans* genetic variants that we investigate here explain a moderate amount of methylation variance (Supplementary Figure 3), the VMR may be influenced by multi-locus genetic effects that explain a greater portion of variance. Furthermore, the LY6S-VMR is an example of an hvCpG cluster and putative human metastable epiallele, suggesting stochastic establishment of DNAm with potential sensitivity to the periconceptional environment^10,11^. Finally, although we did not identify *trans-*mQTL in the KZFP *ZNF525* which also binds the MER11C element in *LY6S-AS1*, we cannot rule out that this also influences epigenetic silencing at this locus. Further work is required to assess the influence of these additional genetic and/or environmental factors on LY6S-VMR methylation.

The function of the *LY6S-AS1* antisense transcript has yet to be defined. It overlaps *LY6S*, a recently annotated interferon-inducible protein-coding gene that is linked to viral resistance and is highly-expressed in a non-classical spleen cell population^24^. Interestingly, there is an IFN-stimulated response element ∼500 bp upstream of *LY6S* that appears in most type I IFN-stimulated genes^24^. Type I IFNs, through their role in fighting viral infection, have been implicated in the early stages of T1D autoimmunity^25^. This suggests a potential link between ZNF808-mediated epigenetic silencing of MER11C, *LY6S-AS1* and aberrant insulin metabolism. Whereas complete *ZNF808* inactivation leads to pancreatic agenesis and neonatal diabetes, we speculate that variable DNA methylation and/or H3K9me3 at the LY6S-VMR could influence the regulation of *LY6S* expression through the action of the *LY6S-AS1* transcript, and that this may be impacted by viral infection leading to increased risk of T1D in severe cases. Additionally, small changes in *LY6S-AS1* transcript levels due to differential methylation states established at the LY6S-VMR could have a small negative impact on pancreas development through epigenetic mosaicism, leading to depleted beta cell numbers with consequent heightened susceptibility to T1D later in life.

The findings we present here provide an example of a primate-specific regulatory network where epigenetic modifications in the early embryo may influence later health. Future investigation of similar candidate loci will establish whether the epigenetic silencing mechanism we describe is linked to other diseases with a developmental origin.

## Methods

### hvCpGs

4,143 hvCpGs representing the top 5% of methylation variance across multiple tissues and ethnicities were identified in a study of 30 publicly available Illumina450K datasets covering 19 unique tissue/cell types and 8 ethnicities. Loci are available in Supplementary Table 5 from Derakhshan et al^11^.

Array background CpGs comprised a set of 406,306 CpGs on the Illumina 450k array covered in at least 15 of the 30 datasets analysed in Derakhshan et al.

### Gambian data

We analysed genomic and methylation data from participants in two studies conducted in the West Kiang region of The Gambia: The Early Nutrition and Immune Development (ENID) trial (ISRCTN49285450)^18,19^ and the Hormonal and Epigenetic Regulators of Growth (HERO-G) study^26^.

#### Methylation data

##### ENID cohort Agilent methyl-seq data

Methylation data for the ENID cohort from DNA isolated from whole blood at age 2 years were previously generated using the Agilent SureSelect Methyl-Seq targeted capture system, focusing on putative metastable epialleles (MEs) and their flanking regions identified in previous studies^16,19,20^. Details on sequencing, alignment and methylation calling are provided elsewhere^19^.

For this study, we analysed CpG methylation in the LY6S-VMR (hg19; chr8: 144120106-144120706). 13 CpGs (out of 28 CpGs in the hg19 reference genome) met our selection criteria of a minimum 30x read depth in at least 300 individuals (Supplementary Table 3). Selected CpGs were covered at a median read depth of 58x (IQR = [44,76]) in a median 344 [332,339] individuals.

##### HERO-G cohort Nanopore methyl-seq data

We generated long-read methyl-seq data for n=177 one-year old infants from the HERO-G cohort on the Oxford Nanopore Technologies (ONT) PromethION 24 platform at MRC Unit The Gambia using R9 (n=71) and R10 (n=106) flow cells with one sample per flow cell. Libraries were prepared from peripheral blood DNA with a target N50 of 10 kb. Reads with a quality score (Qscore) below 9 were removed. Basecalling was performed using Dorado (v0.7.2) using the high accuracy model (dna_r10.4.1_e8.2_400bps_hac@v5.0.0). The resulting reads were aligned to the hg38 reference genome and variant calling was performed using the wf-human_variation pipeline (v2.4.0) using a minimum mean coverage threshold of 20x (n=168 samples). Strand-collapsed methylation calls were extracted using the modkit function in wf-human_variation with modification model 5mC_5hmC (v3). For data visualisation we focussed on all 160 CpGs spanning the LY6S-VMR and MER11C RT + 2 kb either side. Mean [IQR] coverage for CpGs in this region was 23 [15,30], and selected 88 samples with at least 75% of the 160 CpGs covered at a minimum depth of 20x.

#### Genotype data

Genotype data for 445 individuals from the ENID cohort were generated using the Illumina H3Africa array which covers common genetic variants identified in multiple African populations. Genotypes for 2,263,335 SNPs were called using the ‘gtc2vcf’ pipeline (https://github.com/freeseek/gtc2vcf). We excluded SNPs with a GenTrain score below 0.7 and samples with more than 5% missingness, resulting in data for 429 individuals. These genotypes were imputed using SHAPEIT v4.2.2 and IMPUTE5 v1.1.5 with the 1000 Genomes Phase 3 panel as a reference. SNPs with an IMPUTE5 info metric below 0.9 were removed, leaving a total of 47.5M variants, genome-wide.

### Methylation quantitative trait locus mapping using European data from GoDMC

To investigate genetic effects on DNAm at hvCpGs, we used publicly available mQTL data from the Genetics of DNA Methylation Consortium (GoDMC), downloaded from the GoDMC database (http://www.godmc.org.uk/). This provides summary-level data for *cis* and *trans* mQTL at CpGs covered on the Illumina450K array. Significance thresholds of p < 1 x 10^-8^ and p < 1 x 10^-14^ for *cis* and *trans* mQTLs respectively are those used by GoDMC^14^.

### LY6S-VMR methylation quantitative trait locus mapping with Gambian data

For the ENID cohort, we analysed data from a total of 375 individuals with both genotype and methyl-seq data to examine associations between LY6S-VMR methylation and genetic variants in the *ZNF808* gene. The median [IQR] number of samples with sufficient methyl-seq coverage across CpGs in the LY6S-VMR was 344 [332,359].

This analysis focused on *trans*-mQTL at *ZNF808* and its promoter (chr19: 144,120,106 – 144,120,681, hg19); and *cis*-mQTL at the MER11C ChIP-exo peak in the *LY6S-AS1* first intron.

Imputed genetic variants from the Illumina H3Africa array (see above) were filtered based on missingness (< 5%), Hardy-Weinberg equilibrium (p < 0.001), and minor allele frequency (> 1%). This left 78 SNPs in *ZNF808* and a single SNP in the *ZNF808* ChIP-exo peak. mQTL analyses were conducted using an additive linear model with allelic dosage as the predictor and CpG methylation values as the dependent variable, adjusted for sex.

Significant LY6S-VMR – *ZNF808* mQTL associations were defined as those passing a Bonferroni-adjusted significance threshold of 4.9×10^-5^, corresponding to 1,104 tests (78 SNPs x 13 CpGs) at an uncorrected threshold, p = 0.05. We note that this is a conservative threshold given that LY6S-VMR CpGs are highly correlated.

### Retrotransposon coordinates

Coordinates (hg19) for 698 retrotransposon subfamilies belonging to 16 families (including MER11C) were downloaded from the UCSC RepeatMasker database using the Table Browser tool.

### KZFP and TRIM28 binding sites

ChIP-exo data for KZFP and TRIM28 binding sites^1^ from the GEO dataset GSE78099 were obtained using the GEOquery R package (v2.58.0). This dataset includes high-resolution mapping of KZFP binding sites generated in HEK293T cells and endogenous TRIM28 binding sites from H1-ESCs^15^.

### Proximity of hvCpGs to KZFP and TRIM28 binding sites

We used two-sided Fisher’s exact tests to compare the enrichment/depletion of hvCpGs for proximal (<3kb) KZFP and TRIM28 binding sites relative to array background CpGs. We used a de-clustered set of hvCpGs to reduce potential biases due to clustering of hvCpGs^11^. We used a proximity threshold of 3kb because DNAm has been found to spread 3-5kb from the TRIM28/KAP1 docking site in ESCs^27^.

### hvCpG histone modification analysis

H3K modifications in H1-ESCs were examined using the *annotatr* (v1.10.0) R package, using ChIP-seq data generated by the Roadmap Epigenomic Consortium (‘E003’)^28^. We used ‘broadPeak’ data because H3K9me3 histone modifications are diffuse^29^. 3,750 hvCpGs and 318,699 array background CpGs were annotated to these broadPeaks respectively. We tested their enrichment for each type of H3 modification at hvCpGs relative to array background CpGs using two-sided Fisher’s exact tests.

### Transcriptomic and epigenetic regulation in H1 stem cells differentiated toward pancreatic precursors

We reanalysed transcriptomic and epigenetic data published in de Franco et al^13^. We used normalized counts from GEO (GSE205164) and calculated FDR adjusted p-values with DeSEQ2. Epigenetic data was obtained from the same source, mapped to hg19 and the *LY6S-AS1* locus was visualized in IGV.

### insDMR

Coordinates for the insDMR were obtained from Antoun et al^12^, Table 3. The DMR spans 576 bp (chr8:144,120,106 - 144,120,681; hg19) and is located immediately upstream of *LY6-AS1*, previously named *C8orf31* in the hg19 reference.

### Mendelian randomization analysis for investigating causal links

We used two-sample Mendelian randomisation (2SMR) to evaluate the causal relationship between DNA methylation at the LY6S-VMR, and T1D, T2D and pancreas volume (PV). This analysis was conducted using the *TwoSampleMR* R package (v. 0.5.6; https://mrcieu.github.io/TwoSampleMR/) and the MRbase platform^30^, using GWAS summary statistics from the IEU Open GWAS database (https://gwas.mrcieu.ac.uk/datasets/).

We considered methylation at seven CpGs on the Illumina 450k array within the insDMR as our exposure variables (Supplementary Table 3), and eleven associated mQTL (p < 1 x 10^-8^ for *cis* and p < 1 x 10^-14^ for *trans*^14^) from the GoDMC database as genetic instruments (Supplementary Table 9). F-statistics for mQTL instruments ranged from 12.0 to 69.4 (median 31.3) confirming no evidence of weak instrument bias (strong instruments defined as F statistic >10; ^31^).

Summary statistics for these SNPs were extracted from GWAS datasets for T1D, T2D and pancreas volume (Supplementary Table 9). T1D and T2D GWAS were meta-analyses of multiple case-control cohorts (PMIDs and links to papers in Supplementary Table 10).

The following criteria were used for selection of GWAS datasets:

1. Summary statistics available in Open GWAS and therefore accessible via MRbase;
2. European datasets only (since GoDMC mQTL were identified in Europeans);
3. >1M SNPs analysed;
4. Data from a published study with PMID.
5. For T1D, this resulted in 4 GWAS being selected (Supplementary Table 14). We dropped the Sakoue et al. GWAS since the two meta-analysed cohorts in this study (UKBB and FinnGen) were also analysed in the Chiou et al study, but the latter had more cohorts and cases (8 vs 2 cohorts and 18,942 vs 6,447 cases) and used a stricter definition for T1D (see Supplementary Table 14 for further details). For T2D and PV we used all GWAS selected using the above criteria (n=8 and n=1 respectively).

Genetic instruments that were not present in these GWAS were substituted with proxy variants in linkage disequilibrium (r2 > 0.8) from the European phase3 1000G reference panel using MRbase. The effect size for each CpG-outcome pair was determined using the Wald ratio of the percent increase in methylation per allele divided by the unit increase in the outcome per allele. Wald ratios were weighted by the inverse variance of their association with the outcome and combined using the fixed-effects inverse variance weighted method in *TwoSampleMR*.

For the T1D 2SMR, we applied a Bonferroni significance threshold of p = 2.4 x 10^-3^, corresponding to 0.05/21 for 7 CpGs tested across the three analysed GWAS. We note this is a conservative threshold given strong inter-CpGs correlations and given that the three GWAS had a degree of overlapping cases (Supplementary Table 14) and controls.

Additional pleiotropy and causal directionality tests were carried out for the T1D analysis. Horizontal pleiotropy was assessed using the *mr_pleiotropy_test* function in *TwoSampleMR* with Egger_intercept values significantly deviating from zero (p<0.05) considered as evidence of pleiotropy. MR Steiger tests were used to investigate the directionality of causal relationships using the *directionality_test* function in *TwoSampleMR*, with significant tests (p<0.05) supporting a causal effect of insDMR methylation on T1D odds. Results for these additional analyses are presented in Supplementary Table 11 alongside the main MR results.

### Reporting of genomic coordinates

Genomic coordinates in supplementary tables pertaining to GoDMC and ENID mQTL use the hg19 reference as this is used for annotations in the Illumina 450k methylation and H3Africa genotyping array manifests and by the GoDMC mQTL database (http://mqtldb.godmc.org.uk/). Coordinates in Nanopore plots are presented in hg38 as sequences were aligned to that reference.

## Supporting information

Derakhshan_et_al_SUPPLEMENTARY_FIGURES

Derakhshan_et_al_SUPPLEMENTARY_TABLES

## Data Availability

All data produced in the present study are available upon reasonable request to the authors.

## Notes

### Competing Interest Statement

The authors have declared no competing interest.

### Funding Statement

This work was funded by UK Medical Research Council grants MR/ T032863/1 and MC_PC_21037.

### Author Declarations

Ethical approval for all human data analysed in this study was given by the joint Gambia Government/Medical Research Council Unit The Gambia Ethics Committee.

## References

1. Imbeault, M., Helleboid, P. Y. & Trono, D. KRAB zinc-finger proteins contribute to the evolution of gene regulatory networks. Nature 543, 550–554 (2017).

2. Payer, L. M. & Burns, K. H. Transposable elements in human genetic disease. Nat. Rev. Genet. 20, 760–772 (2019).

3. Rosspopoff, O. & Trono, D. Take a walk on the KRAB side. Trends Genet. 39, 844–857 (2023).

4. Gerdes, P., Richardson, S. R., Mager, D. L. & Faulkner, G. J. Transposable elements in the mammalian embryo: pioneers surviving through stealth and service. Genome Biol. 17, 100 (2016).

5. Lawlor, M. A. & Ellison, C. E. Evolutionary dynamics between transposable elements and their host genomes: mechanisms of suppression and escape. Curr. Opin. Genet. Dev. 82, 102092 (2023).

6. Bertozzi, T. M., Elmer, J. L., Macfarlan, T. S. & Ferguson-Smith, A. C. KRAB zinc finger protein diversification drives mammalian interindividual methylation variability. Proc. Natl. Acad. Sci. (2020) doi:10.1073/pnas.2017053117.

7. Costello, K. R. et al. Sequence features of retrotransposons allow for epigenetic variability. eLife 10, e71104 (2021).

8. Bertozzi, T. M. & Ferguson-Smith, A. C. Metastable epialleles and their contribution to epigenetic inheritance in mammals. Semin. Cell Dev. Biol. 97, 93–105 (2020).

9. Morgan, H. D., Sutherland, H. G. E., Martin, D. I. K. & Whitelaw, E. Epigenetic inheritance at the agouti locus in the mouse. Nat. Genet. 23, 314–318 (1999).

10. Derakhshan, M., Kessler, N. J., Hellenthal, G. & Silver, M. J. Metastable epialleles in humans. Trends Genet. 40, 52–68 (2024).

11. Derakhshan, M. et al. Tissue- and ethnicity-independent hypervariable DNA methylation states show evidence of establishment in the early human embryo. Nucleic Acids Res. gkac503 (2022) doi:10.1093/nar/gkac503.

12. Antoun, E. et al. DNA methylation signatures associated with cardiometabolic risk factors in children from India and The Gambia: results from the EMPHASIS study. Clin. Epigenetics 14, 6 (2022).

13. De Franco, E. et al. Primate-specific ZNF808 is essential for pancreatic development in humans. Nat. Genet. 55, 2075–2081 (2023).

14. Min, J. L. et al. Genomic and phenotypic insights from an atlas of genetic effects on DNA methylation. Nat. Genet. 53, 1311–1321 (2021).

15. Nicetto, D. et al. H3K9me3-heterochromatin loss at protein-coding genes enables developmental lineage specification. Science 363, 294–297 (2019).

16. Van Baak, T. E. et al. Epigenetic supersimilarity of monozygotic twin pairs. Genome Biol. 19, 2 (2018).

17. Gunasekara, C. J. et al. A genomic atlas of systemic interindividual epigenetic variation in humans. Genome Biol. 20, 105 (2019).

18. Moore, S. E. et al. A randomized trial to investigate the effects of pre-natal and infant nutritional supplementation on infant immune development in rural Gambia: the ENID trial: Early Nutrition and Immune Development. BMC Pregnancy Childbirth 12, 107 (2012).

19. Candler, T. et al. DNA methylation at a nutritionally sensitive region of the PAX8 gene is associated with thyroid volume and function in Gambian children. Sci. Adv. 7, eabj1561 (2021).

20. Kessler, N. J., Waterland, R. A., Prentice, A. M. & Silver, M. J. Establishment of environmentally sensitive DNA methylation states in the very early human embryo. Sci. Adv. 4, eaat2624 (2018).

21. Juvinao-Quintero, D. L., Sharp, G. C., Sanderson, E. C. M., Relton, C. L. & Elliott, H. R. Investigating causality in the association between DNA methylation and type 2 diabetes using bidirectional two-sample Mendelian randomisation. Diabetologia 66, 1247–1259 (2023).

22. Hancks, D. C. & Kazazian, H. H. Roles for retrotransposon insertions in human disease. Mob. DNA 7, 9 (2016).

23. Deniz, Ö., Frost, J. M. & Branco, M. R. Regulation of transposable elements by DNA modifications. Nat. Rev. Genet. 20, 417–431 (2019).

24. Shmerling, M. et al. LY6S, a New IFN-Inducible Human Member of the Ly6a Subfamily Expressed by Spleen Cells and Associated with Inflammation and Viral Resistance. ImmunoHorizons 6, 253–272 (2022).

25. Newby, B. N. & Mathews, C. E. Type I Interferon Is a Catastrophic Feature of the Diabetic Islet Microenvironment. Front. Endocrinol. 8, (2017).

26. Moore, S. E. et al. Identification of nutritionally modifiable hormonal and epigenetic drivers of positive and negative growth deviance in rural African fetuses and infants: Project protocol and cohort description. Gates Open Res. 4, 25 (2020).

27. Quenneville, S. et al. The KRAB-ZFP/KAP1 system contributes to the early embryonic establishment of site-specific DNA methylation patterns maintained during development. Cell Rep. 2, 766–773 (2012).

28. Kundaje, A. et al. Integrative analysis of 111 reference human epigenomes. Nature 518, 317– 330 (2015).

29. Wang, J., Lunyak, V. V. & Jordan, I. K. BroadPeak: a novel algorithm for identifying broad peaks in diffuse ChIP-seq datasets. Bioinformatics 29, 492–493 (2013).

30. Hemani, G. et al. The MR-Base platform supports systematic causal inference across the human phenome. eLife 7, e34408 (2018).

31. Burgess, S. & Thompson, S. G. Bias in causal estimates from Mendelian randomization studies with weak instruments. Stat. Med. 30, 1312–1323 (2011).

