## Supplementary material for "KZFP-mediated variable DNA methylation of a primate-specific transposon is linked to type I diabetes in humans": Derakhshan_et_al_SUPPLEMENTARY_FIGURES

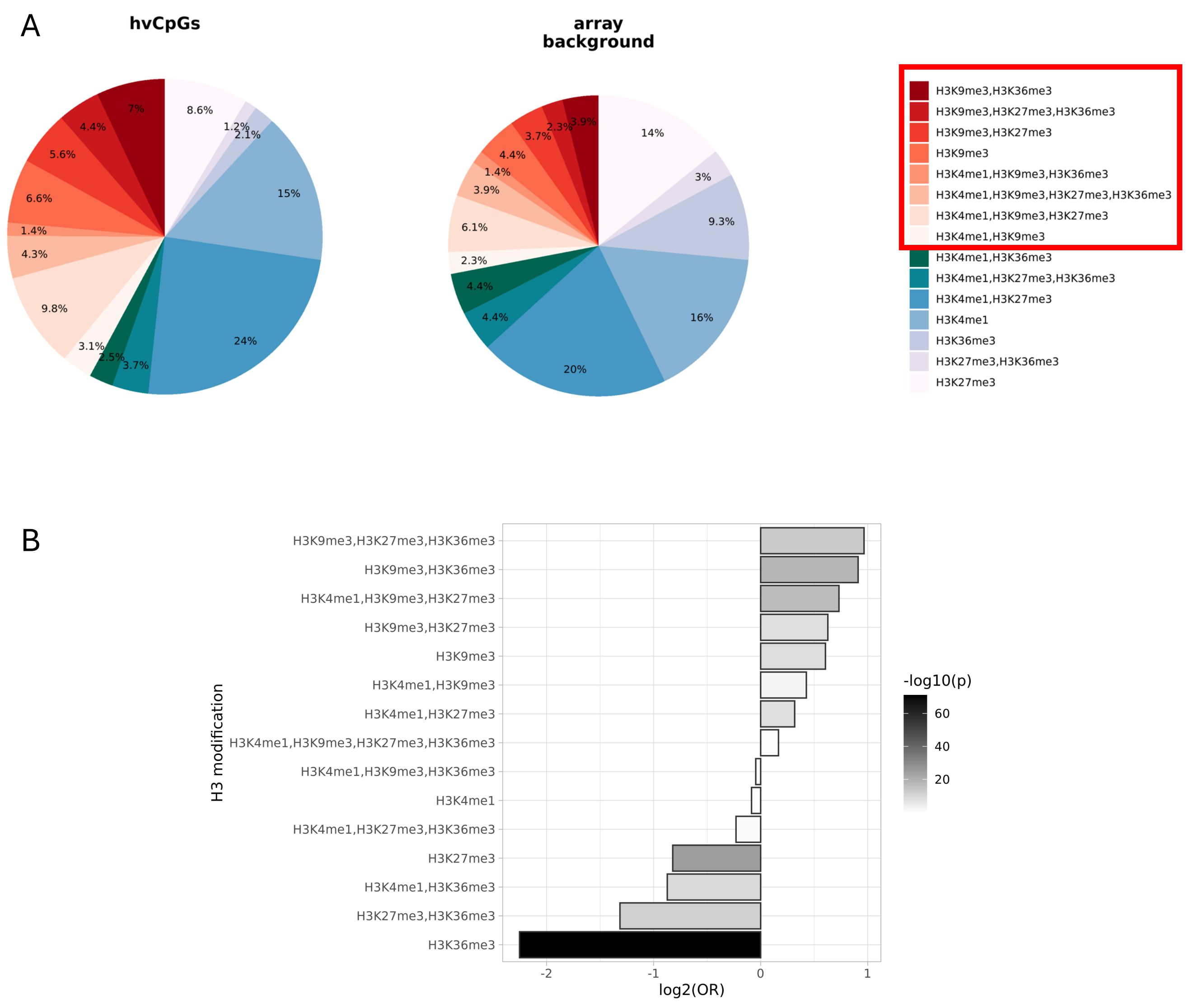


**Supplementary Figure 1. hvCpGs are enriched for proximal H3K9me3 modifications in H1-ESCs.**
**A:** Proportions of 3,750 hvCpGs (left) and 318,699 array background CpGs (right) overlapping each H3 modification signature using 'broadPeaks' ENCODE data obtained in H1-ESCs, identified with the annotatr (v1.10.0) package in R (see Methods). **B:** Odds ratios for enrichment/depletion of hvCpGs overlapping each H3 modification signature relative to Illumina array background. Bar shade gives the Fishers Exact Test (FET) p-values. Only modifications with FDR-adjusted q-values < 0.05 are shown.

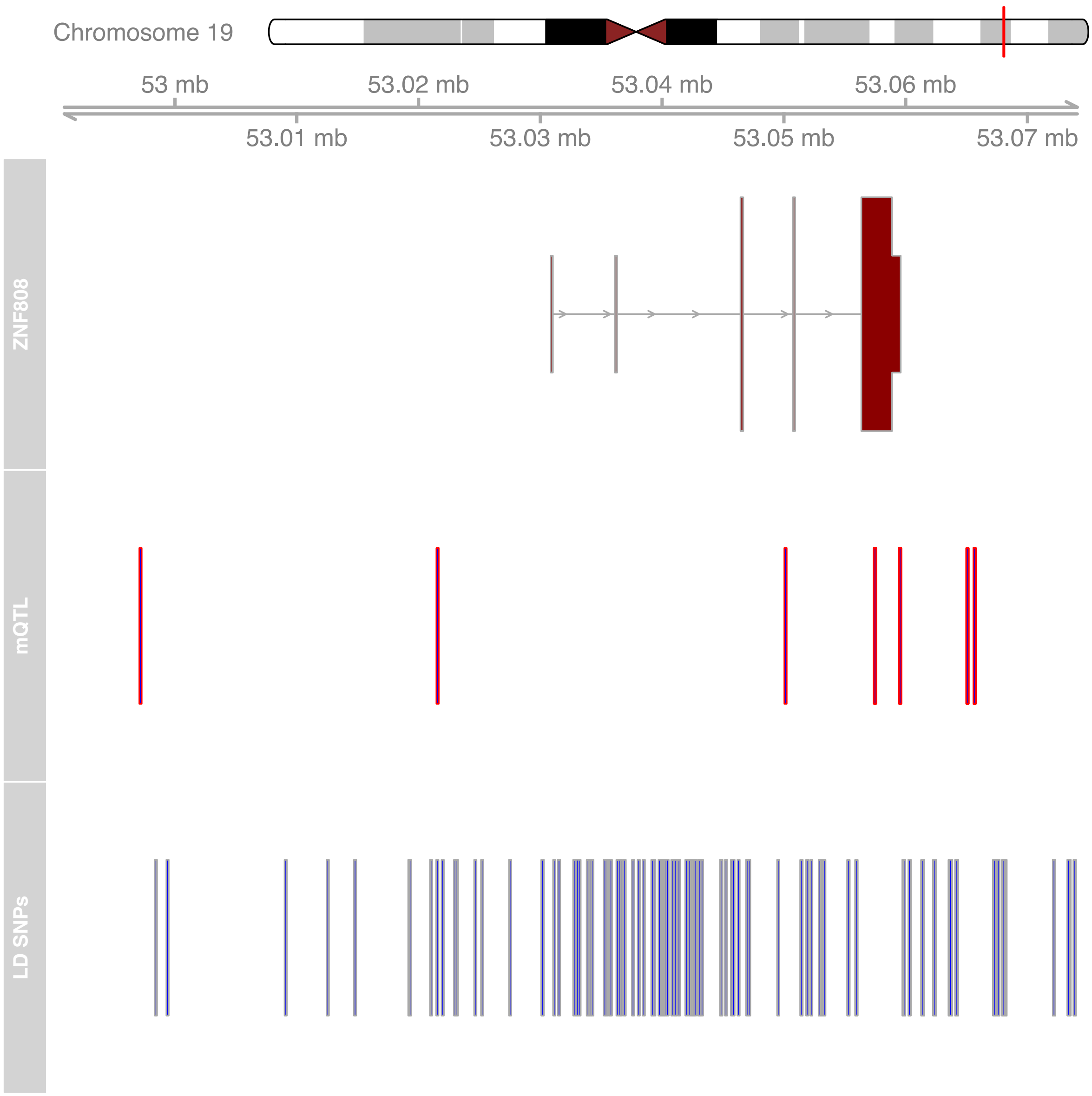


**Supplementary Figure 2. Genomic locations of *ZNF808* trans-mQTL associated with the LY6S-VMR and SNPs in LD.** *ZNF808* track shows the genomic location of the *ZNF808* primary transcript (ENST00000359798). *mQTL* track shows genomic locations of the 7 *ZNF808* trans-mQTL in relation to *ZNF808* (see Supplementary Table 4). *LD SNPs* track shows genomic locations of 116 SNPs in LD ($r^2$>0.8) with identified *ZNF808* mQTL. LD SNPs are identified using the haploR package (v4.07) in R with ldPop='EUR' to specify European ancestry LD mapping.

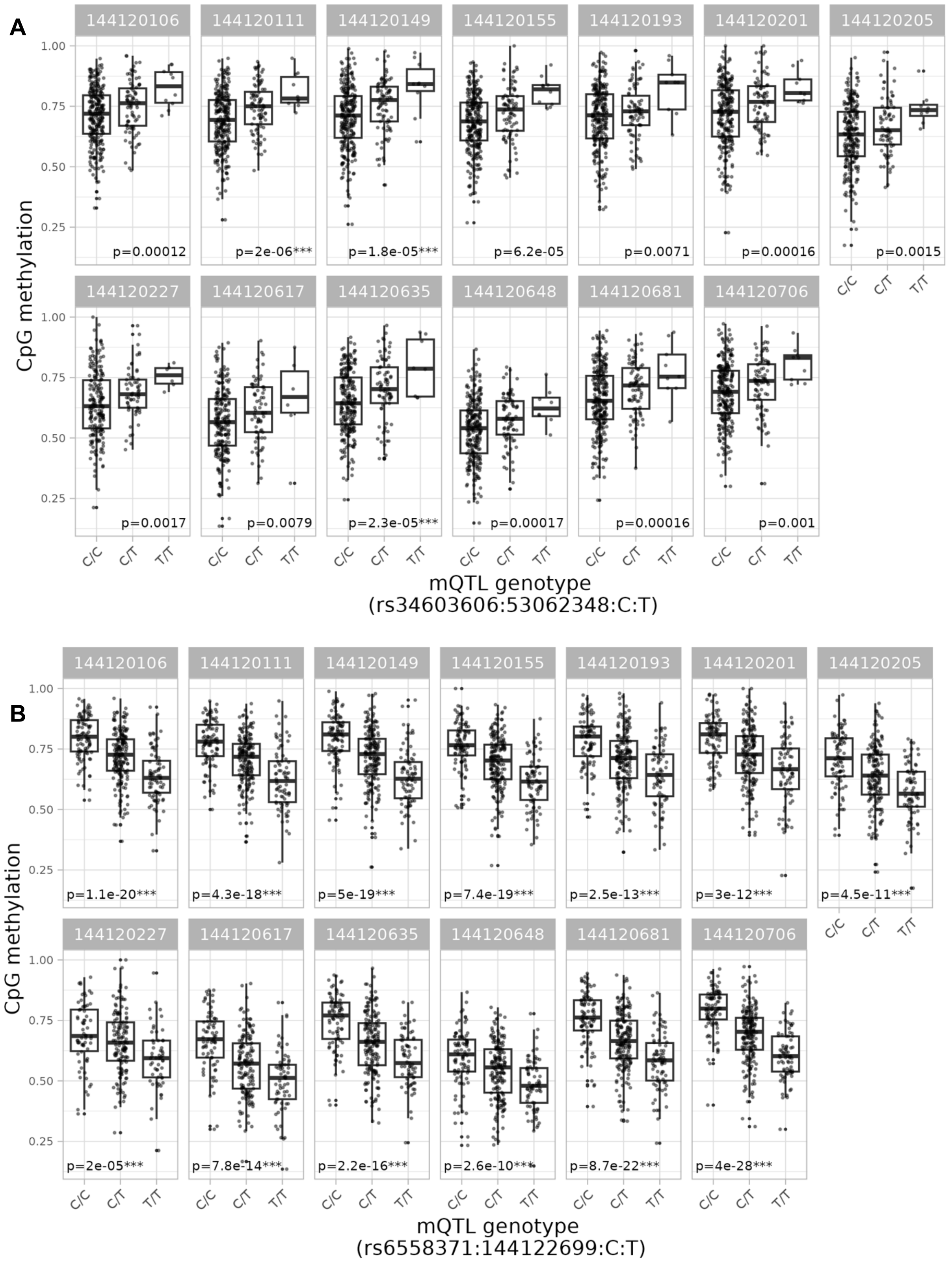


**Supplementary Figure 3. The LY6S-VMR is associated with genetic variation at *ZNF808* in *trans* and its MER11C retrotransposon target in *cis* in data from the Gambian ENID cohort. A:** Box and scatter plots showing methylation distributions at 13 VMR CpGs, stratified by significant *trans*-mQTL genotype (rs34603606:53062348:C:T) in *ZNF808* (see Supplementary Tables 5 & 6). **B:** As C for a single *cis*-mQTL (rs6558371:144122699:C:T) in the ZNF808 CHiP-exo binding site at the proximal MER11C retrotransposon. Individual association p-values for each mQTL-CpG pair are shown. '***' indicates a significant association after Bonferroni correction for multiple tests (see Supplementary Table 6).

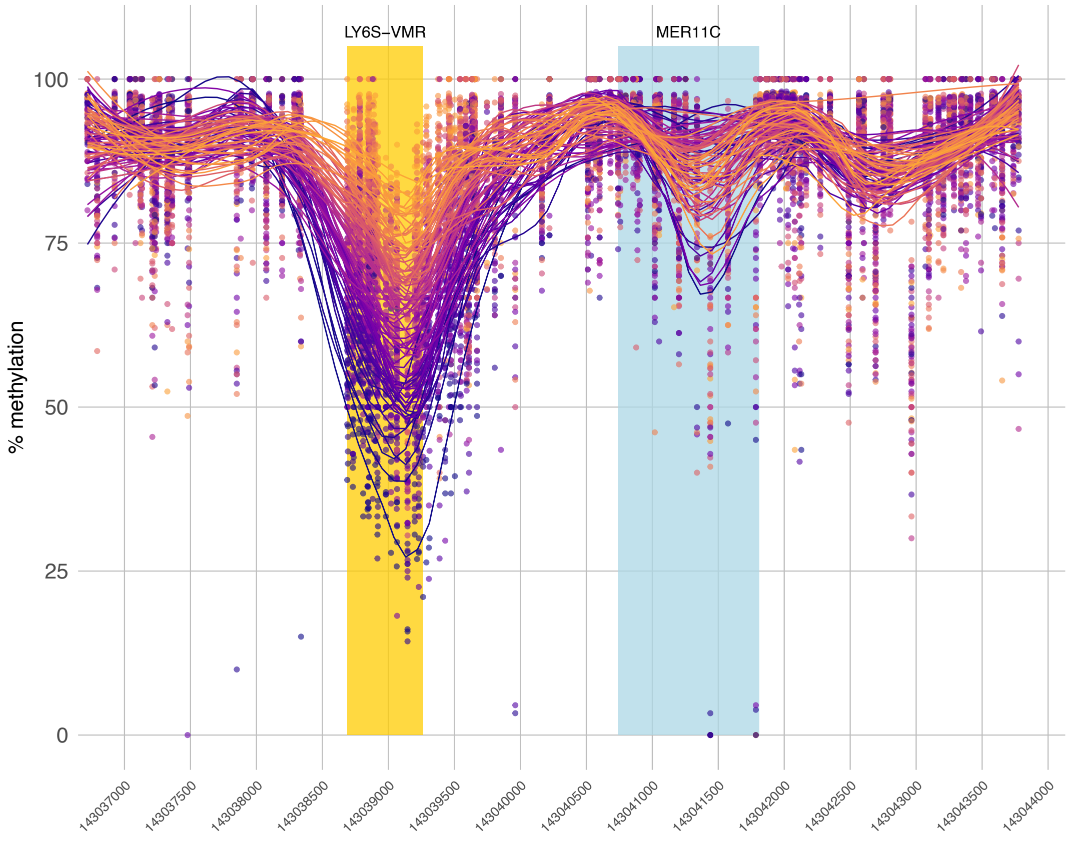


**Supplementary Figure 4. High-resolution characterisation of the LY6S-VMR - MER11C region with Nanopore methyl-seq data.** Plot shows the same data as Figure 3A, but with loess curves summarising regional methylation data for each individual. Colours correspond to ranking by mean methylation across the LY6S-VMR.

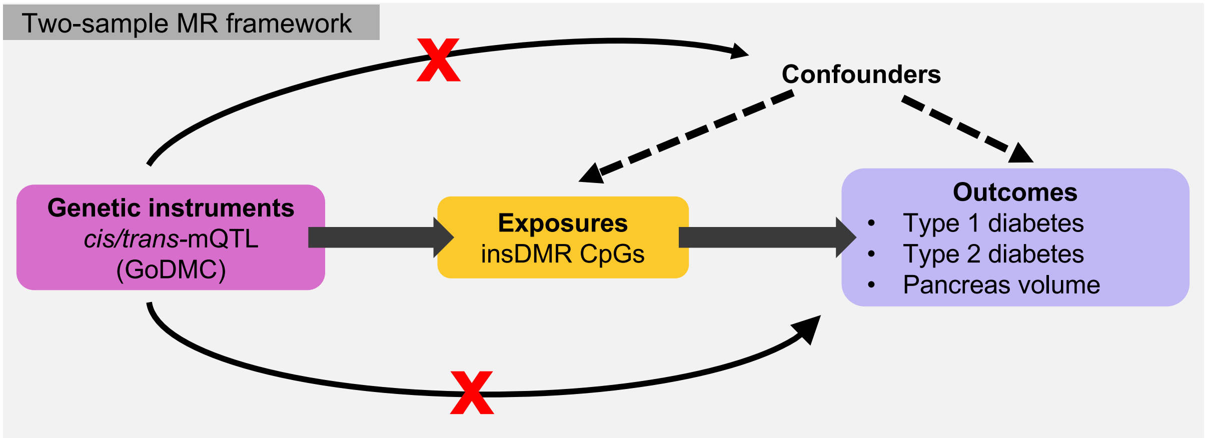


**Supplementary Figure 5. Two-sample Mendelian randomisation (2SMR) framework for investigating a causal link between insDMR CpGs and phenotypes**. Illumina 450k CpGs located in the insDMR are used as 'exposures' and proxied using GoDMC mQTLs ('genetic instruments'). T1D, T2D and pancreas volume are analysed as outcomes. 2SMR assumes that genetic instruments are robustly associated with the exposure, only associated with the outcome via the exposure and not associated with confounders. GoDMC: Genetics of DNA Methylation Consortium (http://mqtldb.godmc.org.uk).

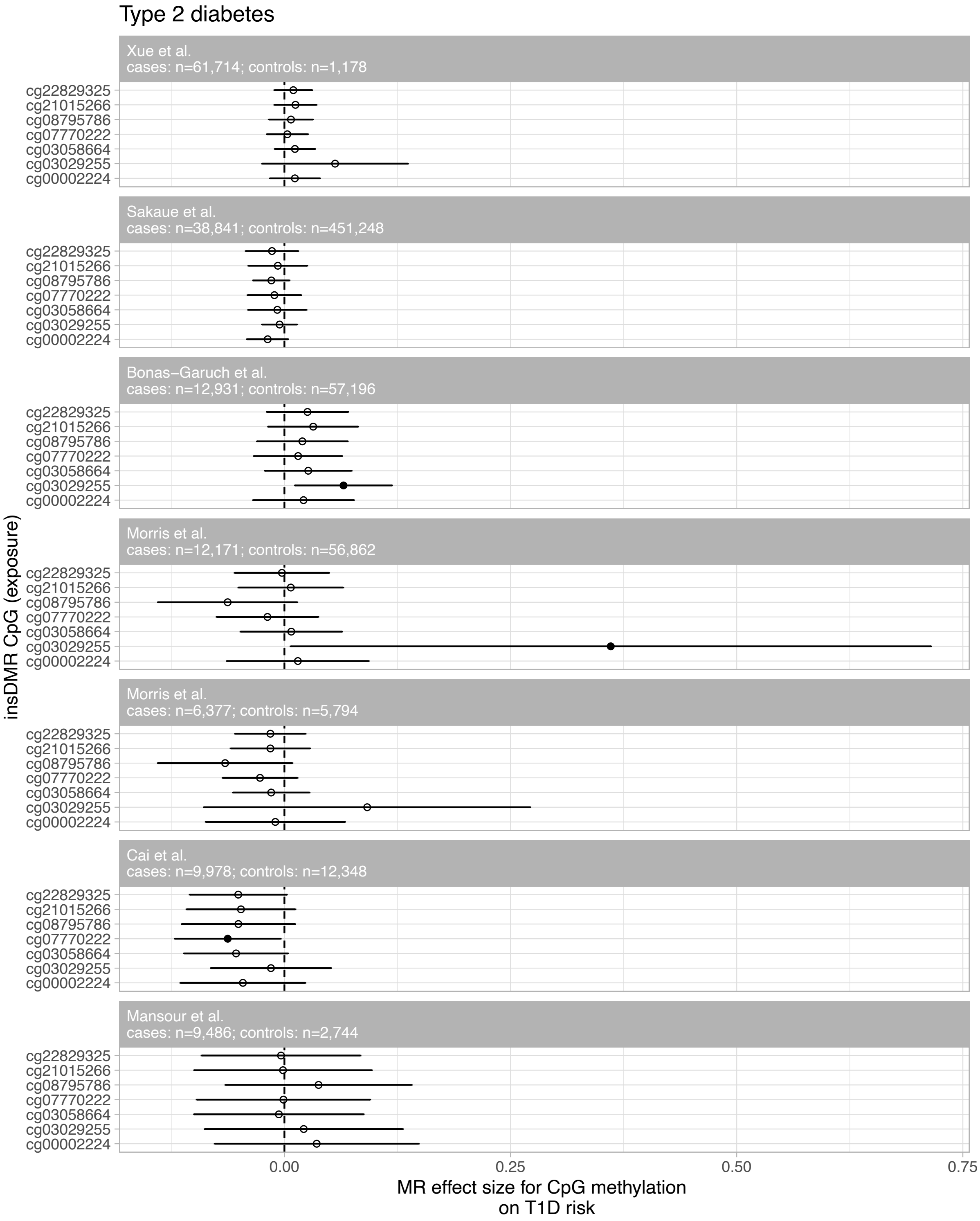


**Supplementary Figure 6. Two-sample Mendelian randomisation analysis of the effect of insDMR methylation on Type 2 diabetes.** See Supplementary Tables 10 & 12 and Methods for further details.

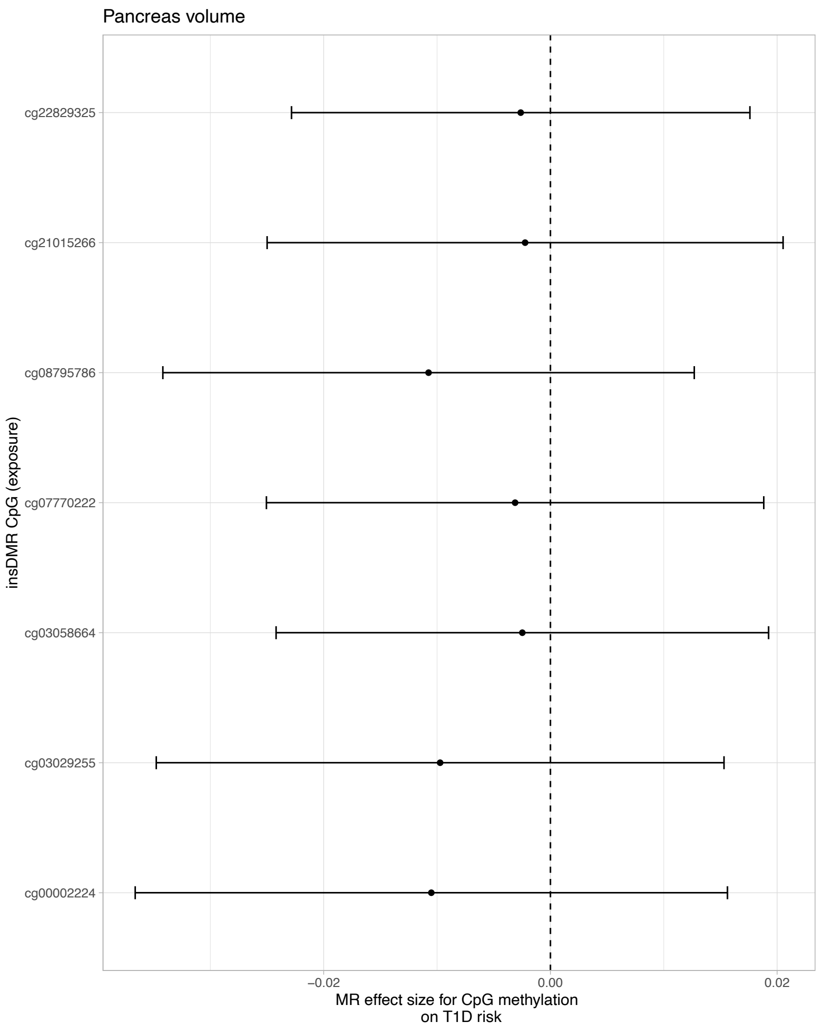


**Supplementary Figure 7. Two-sample Mendelian randomisation analysis of the effect of insDMR methylation on pancreas volume.** See Supplementary Tables 10 and 13 and Methods for further details.
